# Creation of a Clinical Decision-Support Tool for Assigning Occupational Disability to United States Air Force Personnel

**DOI:** 10.1101/2020.05.07.20090530

**Authors:** Maj Colby C. Uptegraft, Catherine T. Witkop

## Abstract

Occupational dispositions (profiles) are the top reason active duty service members are not medically ready to deploy or fulfill their job responsibilities. An audit across multiple U.S. Air Force (AF) medical treatment facilities revealed significant shortcomings in how medical providers assign profiles. We aimed to create a predictive model and a decision-support tool that estimates profile duration.

Using retrospective profiles (n=1,546,805) from the Aeromedical Services Information Management System between 1 Feb 2007 and 31 Jan 2017, we built and validated a decision-support tool that estimates profile length. Multivariate quantile regressions (n=2,575) were performed across five quantiles and six levels of diagnostic specificity for every diagnostic code with 2,100 or more observations.

The models universally estimated profile duration with very poor accuracy *(pseudo*R^2^ 0.000 to 0.168); however, predictive ability was directly correlated with quantile level with minimal variation by diagnostic specificity. Age, O4 to O6+ ranks, very heavy job class, and co-morbid conditions were all significant in more than 25.0% of regressions down all levels of diagnostic specificity. Age, co-morbid conditions, E7-E9 ranks, O4 to O6+ ranks, and light job class all added days to profile duration while E1 to E4 ranks, heavy, and very heavy job class subtracted days.

While this study failed to produce an accurate tool, several findings, the indirect correlation between profile duration and very heavy job class and the assignment of durations based on convenient calendar times, warrant further investigation. For now, providers may consult existing decision-support tools when building profiles for AF service members, heeding attention that they were built with non-representative civilian populations.

**Disclaimer:** The views expressed are solely those of the authors and do not reflect the official policy or position of the US Army, US Navy, US Air Force, the Department of Defense, or the US Government.

## Introduction

Medical readiness—the ability of military members to safely execute their occupational requirements— is the core mission of the Military Health System.^[1,2]^ As of March 2019, 86.8% of Air Force (AF) service members were medically ready.^[3]^ However, to align itself with recent Department of Defense (DoD) retention policy,^[4]^ the AF increased its medical readiness goal to 95%, making readiness improvements a strategic priority.^[5]^

Medical readiness consists of six elements—Periodic Health Assessment, Deployment-(or Duty-) Limiting Conditions (DLCs), Dental Assessment, Immunization Status, Laboratory Studies, and Individual Medical Equipment.^[6]^ Of these DLCs are the primary reason why service members are not medically ready.^[7-9]^ Assigning a DLC is analogous to an occupational disposition in civilian practice; after diagnosing the patient, providers must decide if the ailment interferes with the patient’s ability to fulfill his/her job requirements or deploy.

AF healthcare providers annotate DLCs electronically as a ‘profile’ in the Aeromedical Services Information Management System (ASIMS). An AF profile consists of three categories of restrictions—duty, fitness, and mobility restrictions—the final category being the AF equivalent to DoD DLCs. Each category requires the provider to apply the service member’s medical diagnosis to their in-garrison job, encoded as AF Specialty Codes (AFSCs), AF-wide fitness requirements, and deployment minimums, respectively.

Unfortunately, prior studies show poor agreement between providers and independent subject-matter experts for both the presence and content of these restrictions within profiles [κ = 0.152; 95% CI: 2.226-41.209]. Profiles were both present and within the standard of care only 47% of the time.^[10]^ Under-profiling places individuals in medically compromising work conditions, fitness evaluations, and deployments where access to healthcare services could be limited. Over-profiling may result in underperformance and physical deconditioning, potentially leading to long-term career impacts and negative health implications. Both lead to inaccurate readiness projections on true force availability.

Multiple potential reasons for profiling failure exist. The military depends on its providers to assign occupational dispositions. However, associating diagnoses with a patient’s occupation is not a focus of most medical residencies and fellowships, and non-flight-surgeon providers never receive any formal occupational training. Current AF health information systems also offer minimal decision support to assist with profile creation, thereby allowing a significant potential contribution from human error. While several commercial decision-support solutions exist, namely the Official Disability Guide^®^ (MCG Health, Seattle, WA, USA) and MDGuidelines^®^ (Reed Group, Westminster, CO, USA), both employ propriety algorithms and primarily use disability cases and workers’ compensation claims as data sources.^[11]^ As military populations are generally younger, healthier and have different occupational requirements than civilians, these data sources are not representative of the military, limiting their utility for military practitioners.

Fortunately, when AF medical providers create profiles, ASIMS collects similar data points to elements of these proprietary models. Each profile contains or links to data that includes International Classification of Diseases (ICD) code, age, gender, military rank, AFSC, special duty status, and other co-morbid medical conditions. Thus, an extensive, AF-specific dataset is available to create a similar predictive tool using a representative population.

### Objective

The purpose of this study is to create a predictive model and decision-support tool for profile duration using a quantile regression approach and AF profiling data from ASIMS. A profile decision-support tool could offer improved point-of-care information to medical providers and ultimately improve the validity of readiness metrics.

## Methods

### Overview

Data for this study were supplied by ASIMS for all finalized profiles between 1 Feb 2007 and 31 Jan 2017. These data included all profiles (n=1,546,805) that were signed by a healthcare provider and/or profile officer. Exclusion criteria were restrictions associated with V00-Z99 and DoD ICD-10-CM codes and profiles for pregnant service members (assignment availability code 81s), service members concurrently undergoing a medical evaluation board (assignment availability code 37s), or service members with permanent mobility restrictions (any assignment limitation codes). All analyses were performed with Stata/IC 14 for Windows. This study was reviewed and approved as exempt by the Uniformed Services University Institutional Review Board (FWA 00001628).

### ICD Mapping

Primary diagnosis ICD-9-CM codes were mapped to ICD-10-CM codes using the 2017 Centers for Medicare & Medicaid Services’ General Equivalence Mapping documentation.^[12]^ For ICD-9-CM codes that matched to multiple ICD-10-CM codes, only the ICD-10-CM code that had the greatest count in the ICD-10-CM-only subset (Profiles published after 30 Sep 2015) was kept and considered the one-to-one match.^[13]^ The remaining matches were discarded. Observations with primary diagnoses that failed conversion (n=6,325) and all V00-Z99 and DoD ICD-10-CM codes (n=1,218) were dropped. Additionally, all observations with a primary diagnosis of duration less than one day or greater than 364 days were dropped (n=38,693).

ICD-10-CM code morbidity mapping documentation was obtained from the Armed Forces Health Surveillance Branch.^[14]^ All profile diagnoses—primary, secondary, and tertiary—were mapped to one of 25 morbidity categories and one of 143 morbidity subcategories.

### Conversion of Air Force Specialty Codes to Job Classes

Officer and enlisted AFSCs were extracted from their respective classification directories.^[15,16]^ These AFSCs were crosswalked to Standard Occupational Classifications using the Military Occupational Classification crosswalking module provided by the Occupational Information Network.^[17]^ Each Standard Occupational Classification was then crosswalked to an equivalent occupation listed in the Dictionary of Occupational Titles, Fourth Edition.^[18]^ In the cases where multiple Standard Occupational Classifications matched to one AFSC and/or where multiple occupations in the Dictionary of Occupational Titles matched to one Standard Occupational Classification, the best one-to-one match for each crosswalk was decided by the author. This occupational mapping resulted in each AFSC matched one-to-one with an occupation listed in the Dictionary of Occupational Titles.

Each occupation listed in the Dictionary of Occupational Titles has a respective strength rating (job class) of sedentary, light, medium, heavy, or very heavy work.^[19]^ Job classes of each of the occupations were then assigned to the matching AFSC. Job classes for AFSCs unable to be mapped to standard occupations were decided by the author. Enlisted AFSCs with a ‘9’ as the fourth digit and officer codes with a ‘4’ as the fourth digit signify a more administrative, and less physically active, occupation compared to other AFSCs of the same category. Thus, these respective job classes were reduced by one ordinal level if the strength rating assigned was not the lowest ordinal level, sedentary work. All ranks of colonel and above (O6+) were assigned a ‘sedentary work’ job class.

Specialty duty status for AFSCs was determined through the AF Enlisted and Officer Classification Directories.^[15,16]^ AFSCs were assigned a ‘Yes’ for specialty duty status if the code required any type of ground-based controller, flying class, and/or personnel reliability program qualification prior to entry into the career field.

### Model Development

To estimate profile duration by ICD-10-CM diagnosis, multivariate quantile regressions with pairs-bootstrapping (100 repetitions) were performed across five quantiles and down six levels of diagnostic specificity. The five quantiles were 0.10, 0.25, 0.50 (Median), 0.75, and 0.90. The first five levels of specificity were all possible lengths of ICD-10-CM codes, from three to seven characters, of the primary diagnosis of each profile. For instance, at the fifth level, the maximum allowable ICD length was three characters; thus, the codes A06.01 and A06.02 would both be A06 at this level. The sixth level of diagnostic specificity was the morbidity category of the primary diagnosis of each profile.

A quantile regression was performed at each level of diagnostic specificity and across the five quantiles for all ICD-10-CM codes / morbidity subcategories with 2,100 or more observations. This approach resulted in 2,575 total regressions. A minimum observation count of 2,100 was selected as the estimated sample size needed to obtain a power of 0.80 with 14 total covariates and to detect a small effect size (f^2^ = 0.02).^[20,21]^ Multiple diagnostic levels were included to increase the available sample size and diagnostic range and thus include more diagnoses in the predictive tool (**Figure 1**). However, the minimum observation count of 2,100 excluded 128,391 (8.6%) observations from regression analysis.

**Figure 1.**
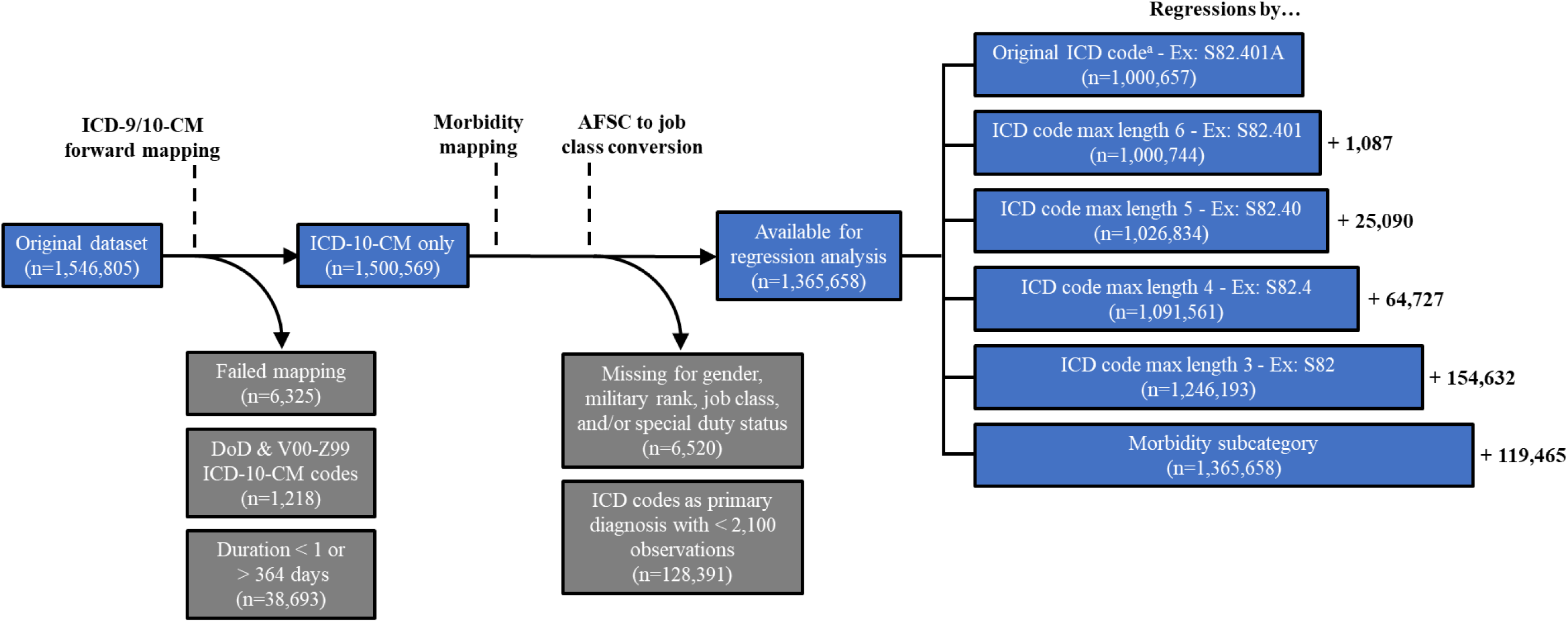
Data cleaning, mapping, and regression allocation process ^a^Independent of ICD code length, by the original ICD code in the profile AFSC, Air Force Specially Code;DoD, Department of Defense; ICD-9/10-CM, International Classification ofDiseases, Ninth/Tenth Revision, Clinical Modification.

The dependent variable was the duration of the primary diagnosis in days while the independent variables included age, gender, military rank, job class, special duty status, co-morbid conditions, and similar co-morbid conditions. Co-morbid conditions were defined as the presence of any secondary or tertiary diagnosis in the profile. Similar co-morbid conditions were defined as the presence of any secondary or tertiary diagnosis within the same morbidity subcategory as the primary diagnosis.

Variance inflation factors for independent variables were all less than 10; thus, it was concluded that collinearity among variables was not significantly present. However, the mean variance inflation factor was 2.3, which might be considered significantly greater than 1.0, making multicollinearity a concern.^[22]^ The highest variance inflation factors were observed among age and the lower enlisted rank groups (Age: 3.3; E1-E4: 6.6; E5-E6: 5.6).

All observations missing or masked for either gender, military rank, job class, and/or specialty duty status (n=6,520 [0.4%]) were excluded from the quantile regression analysis as these variables will be known at the point of care, yielding a final sample size of 1,365,658 profiles (**Table I**).

**Table I.** Demographics of U.S. Air Force profiles used in regression analysis, 1 Feb 2007 - 31 Jan 2017 (n=1,365,658)

|  |  |  |
| --- | --- | --- |
| Mean age, years (sd) |  | 29.5 (7.3) |
| Age group | 25 or younger | 499,205 (36.6) |
|  | 26 - 30 | 327,687 (24.0) |
|  | 31 - 35 | 233,722 (17.1) |
|  | 36 - 40 | 188,463 (13.8) |
|  | 41+ | 116,581 (8.5) |
| Gender | Female | 332,529 (24.3) |
|  | Male | 1,033,129 (75.7) |
| Military rank group | E1 - E4 | 538,256 (39.4) |
|  | E5 - E6 | 538,321 (39.4) |
|  | E7 - E9 | 148,404 (10.9) |
|  | O1 - O3 | 69,207 (5.1) |
|  | O4 - O5 | 60,122 (4.4) |
|  | O6+ | 11,348 (0.8) |
| Job class | Sedentary | 331,367 (24.3) |
|  | Light | 438,327 (32.1) |
|  | Medium | 312,113 (22.9) |
|  | Heavy | 237,285 (17.4) |
|  | Very Heavy | 46,566 (3.4) |
| Special duty status | No | 1,242,668 (91.0) |
|  | Yes | 122,990 (9.0) |
| Co-morbid conditions | No | 1,217,205 (89.1) |
|  | Yes | 148,453 (10.9) |
| Similar co-morbid conditions | No | 1,360,496 (99.6) |
|  | Yes | 5,162 (0.4) |
| **Total** |  | **1,365,658** |

Note: Data are n (%) unless otherwise noted.
E1, Airman Basic; E4, Senior Airman; E7, Master Sergeant; E9, Chief Master Sergeant; O1, Second Lieutenant; O3, Captain; O4, Major; O5, Lieutenant Colonel; O6+, Colonel and above.

### Model Validation

k-fold cross-validation with 10 subsamples was performed on each quantile regression to assess the model fit with out-of-sample observations. These cross-validation quantile regressions were performed without bootstrapping and under the assumption that the model variables were independent and identically distributed to reduce computational time. Since bootstrapping does not affect model fit, this approach did not affect the *pseudo*R*^2^* values for the cross-validation subsamples.^[23]^ The *pseudo*R*^2^* of the 10 subsamples for each regression were averaged to determine the overall validated fit.

### Web Application Design

The decision-support tool was built with R and RShiny and is available at <https://colbycoapps.shinyapps.io/mrpt/>.

## Results

### Overall Fit & Validation

The fit and validation statistics for all categories and individual regressions were very poor with the *pseudo*R^2^ ranging from 0.000 to 0.168. Thus, at best the model covariates account for 16.8% of the variance in profile duration and some models failed completely.

However, lower levels of diagnostic specificity exhibited similar model fit (mean *pseudo*R^2^) and model validation (k-fold cross-validation *pseudo*R^2^ mean) as higher levels of specificity (**Table II**). Quantile regressions at the maximum three-character level and morbidity subcategory level of specificity resulted in similar, albeit poor, fit and validation statistics as higher levels of specificity.

**Table II.**
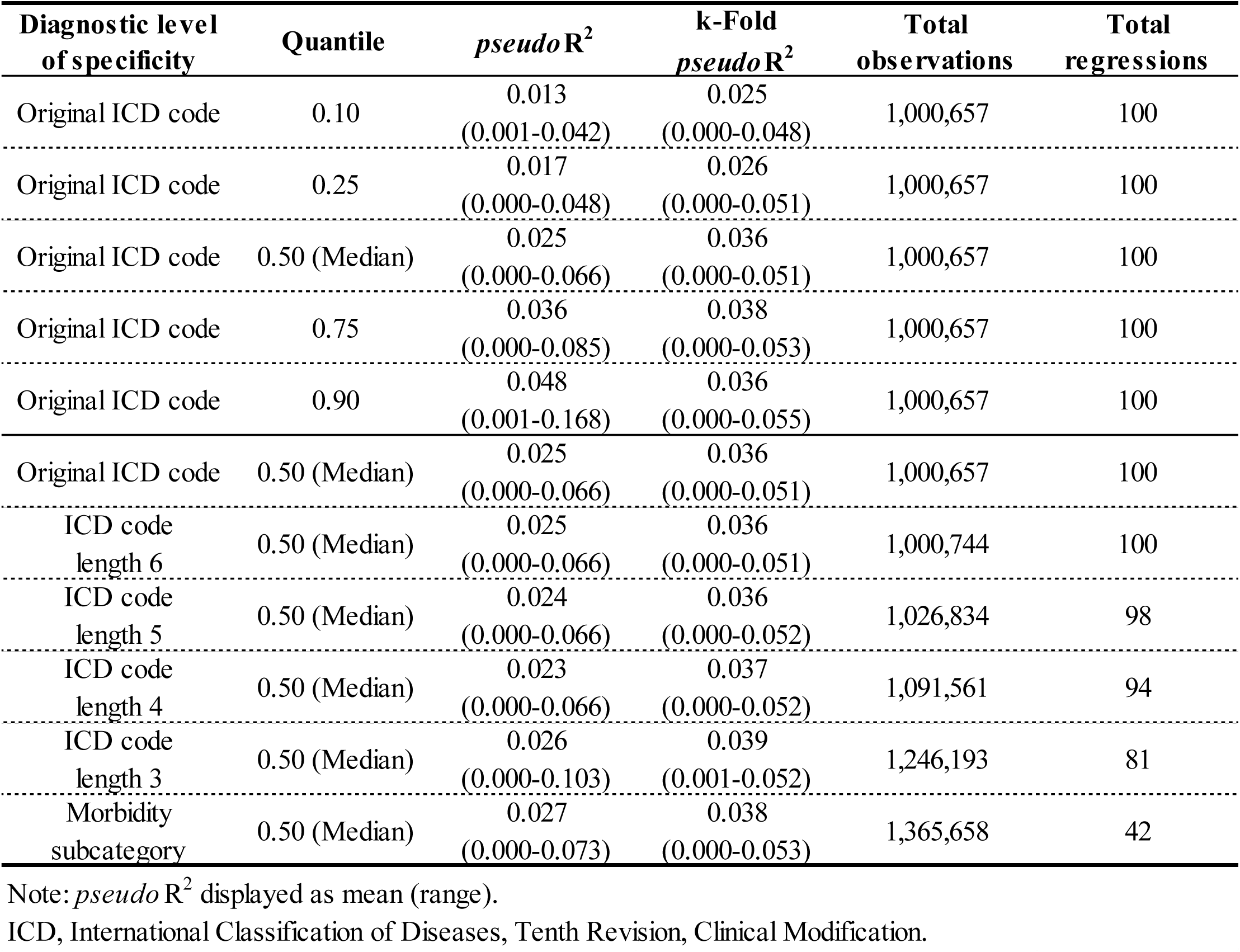
Regression fit & validation statistics [sample], by diagnostic level of specificity & quantile

| Diagnostic level of specificity | Quantile | *pseudo R*<sup>2</sup> | k-Fold *pseudo R*<sup>2</sup> | Total observations | Total regressions |
| --- | --- | --- | --- | --- | --- |
| Original ICD code | 0.10 | 0.013<br>(0.001-0.042) | 0.025<br>(0.000-0.048) | 1,000,657 | 100 |
| Original ICD code | 0.25 | 0.017<br>(0.000-0.048) | 0.026<br>(0.000-0.051) | 1,000,657 | 100 |
| Original ICD code | 0.50 (Median) | 0.025<br>(0.000-0.066) | 0.036<br>(0.000-0.051) | 1,000,657 | 100 |
| Original ICD code | 0.75 | 0.036<br>(0.000-0.085) | 0.038<br>(0.000-0.053) | 1,000,657 | 100 |
| Original ICD code | 0.90 | 0.048<br>(0.001-0.168) | 0.036<br>(0.000-0.055) | 1,000,657 | 100 |
| Original ICD code | 0.50 (Median) | 0.025<br>(0.000-0.066) | 0.036<br>(0.000-0.051) | 1,000,657 | 100 |
| ICD code length 6 | 0.50 (Median) | 0.025<br>(0.000-0.066) | 0.036<br>(0.000-0.051) | 1,000,744 | 100 |
| ICD code length 5 | 0.50 (Median) | 0.024<br>(0.000-0.066) | 0.036<br>(0.000-0.052) | 1,026,834 | 98 |
| ICD code length 4 | 0.50 (Median) | 0.023<br>(0.000-0.066) | 0.037<br>(0.000-0.052) | 1,091,561 | 94 |
| ICD code length 3 | 0.50 (Median) | 0.026<br>(0.000-0.103) | 0.039<br>(0.001-0.052) | 1,246,193 | 81 |
| Morbidity subcategory | 0.50 (Median) | 0.027<br>(0.000-0.073) | 0.038<br>(0.000-0.053) | 1,365,658 | 42 |

Note: *pseudo R*<sup>2</sup> displayed as mean (range).
ICD, International Classification of Diseases, Tenth Revision, Clinical Modification.

Mean *pseudo*R^2^ values were directly correlated with regression quantile (**Table II**). These values for the original ICD code increased from 0.013 for quantile 0.10 to 0.048 for quantile 0.90, indicating that the model accounts for more variance in profile duration as duration increases.

### General Coefficient Influence on Profile Duration

At quantile 0.50 and when compared to their respective reference category, age, O4-O5 ranks, O6+ ranks, very heavy job class, and co-morbid conditions were all significant in more than 25.0% of regression models down all levels of diagnostic specificity (**Table III**). The presence of co-morbid conditions stood out as being significant in around 80% of models. A similar pattern of significance, with minor deviations, was exhibited across quantiles for regressions by the original ICD code (**Table IV**).

**Table III.** Regression summary statistics, quantile 0.50 (median), by diagnostic level of specificity

|  |  | Original<br>ICD code | ICD code<br>length 6 | ICD code<br>length 5 | ICD code<br>length 4 | ICD code<br>length 3 | Morbidity<br>subcategory |
| --- | --- | --- | --- | --- | --- | --- | --- |
| Total regressions |  | 100 | 100 | 98 | 94 | 81 | 42 |
| Age, years |  | 0.7 (32.6) | 0.7 (33.7) | 0.7 (32.2) | 0.7 (30.4) | 0.6 (38.4) | 0.4 (51.2) |
| Gender | Female |  |  |  | ref |  |  |
|  | Male | 7.0 (27.2) | 11.2 (22.8) | 8.5 (23.3) | 7.9 (22.8) | 4.3 (26.7) | 0.6 (39.0) |
| Military<br>rank<br>group | E1 - E4 | -4.5 (15.2) | -8.3 (15.2) | -8.3 (15.6) | -5.2 (14.1) | -7.1 (17.4) | -5.6 (24.4) |
|  | E5 - E6 | 9.4 (6.5) | 9.4 (6.5) | 12.2 (6.7) | 7.8 (5.4) | -3.3 (3.5) | 3.6 (12.2) |
|  | E7 - E9 | 15.5 (15.2) | 17.3 (15.2) | 15.6 (14.4) | 19.4 (7.6) | 7.1 (9.3) | 7.5 (26.8) |
|  | O1 - O3 |  |  |  | ref |  |  |
|  | O4 - O5 | 15.6 (30.4) | 15.2 (32.6) | 14.2 (28.9) | 14.9 (30.4) | 13.4 (31.4) | 11.1 (36.6) |
|  | O6+ | 27.4 (35.9) | 26.2 (35.9) | 24.8 (38.9) | 25.5 (37.0) | 18.7 (37.2) | 28.0 (43.9) |
| Job<br>class | Sedentary |  |  |  | ref |  |  |
|  | Light | 6.4 (8.7) | 5.7 (9.8) | 5.7 (10.0) | 6.8 (7.6) | 5.2 (7.0) | 1.5 (12.2) |
|  | Medium | 0.1 (6.5) | -0.4 (8.7) | -0.7 (11.1) | -1.2 (7.6) | -3.1 (10.5) | -1.1 (19.5) |
|  | Heavy | -2.6 (8.7) | -2.8 (10.9) | -1.2 (11.1) | -3.7 (13.0) | -4.9 (17.4) | -2.9 (14.6) |
|  | Very Heavy | -8.5 (38.0) | -8.6 (39.1) | -8.7 (41.1) | -9.7 (37.0) | -7.8 (43.0) | -5.5 (51.2) |
| Special<br>duty<br>status | No |  |  |  | ref |  |  |
|  | Yes | 3.6 (10.9) | 6.1 (14.1) | 2.3 (12.2) | -0.6 (12.0) | 2.4 (10.5) | 0.1 (24.4) |
| Co-morbid<br>conditions | No |  |  |  | ref |  |  |
|  | Yes | 19.3 (79.4) | 18.7 (82.6) | 19.0 (80.0) | 18.8 (84.8) | 18.4 (86.1) | 22.5 (82.9) |
| Similar<br>co-morbid<br>conditions | No |  |  |  | ref |  |  |
|  | Yes | -8.1 (12.0) | -8.5 (16.3) | -9.4 (15.6) | -7.4 (17.4) | -3.0 (16.3) | 6.0 (22.0) |

Note: Data are average $\beta$ value (% significant) unless otherwise noted. The average $\beta$ value is the average coefficient value across the total number of regressions. Only statistically significant coefficients ( $p < 0.05$ ) were included in the average. The % significant is the percentage of coefficients that were statistically significant ( $p < 0.05$ ) out of the number of regressions in the 'Total Regressions' column. All ICD codes longer than the indicated maximum were truncated to the meet the maximum character length at each respective level.
E1, Airman Basic; E4, Senior Airman; E7, Master Sergeant; E9, Chief Master Sergeant; O1, Second Lieutenant; O3, Captain; O4, Major; O5, Lieutenant Colonel; O6+, Colonel and above; ICD, International Classification of Diseases, Tenth Revision, Clinical Modification.

**Table IV.**
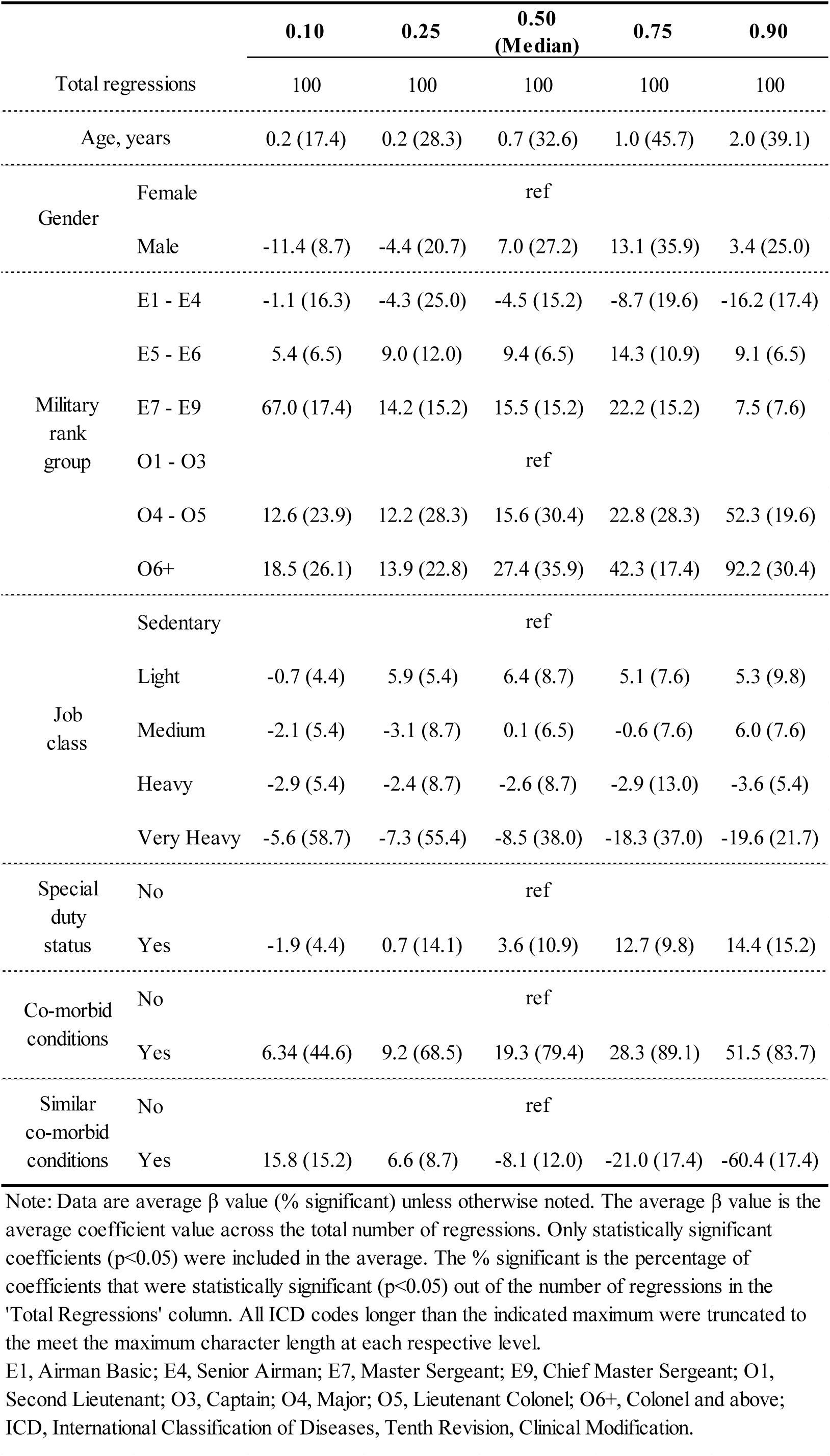
Regression summary statistics, original ICD code, by quantile

### Direction of Coefficient Influence on Profile Duration

When significant (p<0.05) and compared to their respective reference category, age, E7-E9 ranks, O4-O5 ranks, O6+ ranks, and co-morbid conditions all added days to profile duration across all quantiles and down all levels of diagnostic specificity. When significant (p<0.05) and compared to their respective reference category, E1-E4 ranks, heavy job class, and very heavy job class all subtracted days from profile duration across all quantiles and down all levels of diagnostic specificity. The remaining coefficients displayed mixed effects across quantiles and/or diagnostic levels of specificity (**Tables III–IV**).

At lower quantiles (0.10 and 0.25), male gender subtracted days from profile duration while adding days at higher quantiles (0.50, 0.75, and 0.90) when compared to female gender. When compared to O1-O3 ranks, E5-E6 ranks added days to profile duration across all quantiles except at the maximum three-letter level of diagnostic specificity for quantile 0.50. Light job class subtracted days from profile duration at quantile 0.10 but added days to profile duration at all remaining quantiles and diagnostic levels of specificity when compared to sedentary job class. Medium job class mostly detracted days from profile duration except at quantile 0.50 and 0.90 for the original ICD code while special duty status generally added days to profile duration except at quantile 0.10 and the ICD code max length four level of specificity. Lastly, similar co-morbid conditions added days to profile duration at quantiles 0.10 and 0.25 and the morbidity subcategory level of specificity, but subtracted days throughout the remaining quantiles and levels (**Tables III-IV**).

## Discussion

Overall, the mean *pseudo*R^2^ for both model fit and the cross-validations were very poor, indicating that models do not explain much of the variance in profile duration. However, *pseudo*R^2^ values increased as quantiles increased, indicating that the models’ independent covariates can predict profile duration with greater accuracy at higher quantiles than lower quantiles (**Table IV**).

Six profile durations (7, 14, 30, 60, 90, and 180 days) accounted for 45.2% of all observations. This suggests many medical providers assign profile durations based on convenient calendar intervals rather than disease prognosis, patient characteristics, or occupational status, which might explain the models’ poor performance. However, providers might be exhibiting more clinical judgement when assigning profiles of longer duration. Future studies should include the provider assigning the profile as an additional covariate.

Lower levels of diagnostic specificity exhibited similar *pseudo*R^2^ values as higher levels (**Table II**). As specificity decreases, one would expect the predictive ability to decrease as well. However, since providers seemingly assign profiles by calendar convenience, this indiscriminate duration assignment would affect the power to detect smaller effect sizes. Higher levels of specificity had lower sample sizes per regression than lower levels; thus, these higher levels might have been underpowered to detect smaller effects. Therefore, the minimum sample size of 2,100 observations per regression might have been inadequate.

### Independent Covariates

The effects of age, E7-E9 ranks, O4-O5 ranks, O6+ ranks, and co-morbid conditions to add to profile duration are not surprising (**Tables III-IV**). Profile duration would be expected to increase as age increases and if co-morbid conditions are present. Some of the additive effect of these rank categories could either be from collinearity with age or a real effect when compared to the O1-O3 reference category and after controlling for age.

Counterintuitively, heavy and very heavy job classes subtracted days from profile duration on average when compared to sedentary job class (**Tables III-IV**). This finding contradicts prior research of U.S. Army occupational classes and disability using a job classification scheme similar to the Dictionary of Occupational Titles.^[24]^ While heavy job class was significant only a small percentage of the time (5.4% - 17.4%), very heavy job class was among the top-three most significant coefficients across quantiles and levels of specificity (37.0% - 58.7%), making its negative effect on profile duration hard to discount. When mapping AFSCs to job classes, all special forces AFSCs were converted to very heavy job class. These individuals, while likely prone to disability long term, are generally known to avoid profiles, even when experiencing significant pain. While their profile avoidance has not been validated or studied, fear of washing out of training, disappointing their fellow Airmen, missing mission opportunities, and/or differences in perception of what qualifies as an ‘injury’ or pain necessitating rest and recovery might explain this phenomenon. Profile avoidance would also explain attenuated profiles since these members would seek medical clearance faster than others.

One of the advantages of using quantile over linear regression is its ability to detect covariate influences across the entire range of a conditional distribution. Male gender, light job class, medium job class, and special duty status tended to subtract days from profile duration at lower quantiles and add days at higher quantiles (**Table IV**). These differential effects across quantiles are difficult to explain. For gender, males might receive shorter profiles on average than females but may be more likely to suffer serious injuries or medical conditions requiring longer disability.^[25, 26]^ For instance, males were much more likely than females to receive profiles for alcohol abuse or dependence, and the median profile duration for these diagnoses was 177 and 179 days, respectively, much longer than most other conditions (**data not shown**). A similar phenomenon could explain special duty status. These service members might generally receive shorter profiles to return to ‘up’ status faster but are more likely to suffer serious injuries and/or conditions given the nature of their work.

Similar co-morbid conditions displayed the opposite trend, adding days to duration at lower quantiles and subtracting days at higher ones (**Table IV**). Multiple similar conditions might compound a simple problem, such as generic musculoskeletal pain, but generalized complaints at higher quantiles might signify a less serious underlying pathology and therefore a quicker recovery time.

The overall poor fit and validation statistics do not discount the effect of the independent covariates on profile duration. While most covariates were significant in influencing profile duration less than half the time (**Tables III-IV**), using them to predict duration would still be superior than relying on medians or percentiles alone. However, considering the models’ poor performance, medical providers should only use a tool powered by these regressions as a reference point, rather than as an accurate prediction of profile duration.

### Limitations

The accuracy of ICD codes for determining diseases rates or health indices varies. Most studies assert that their accuracy is disease dependent, ranging from 50 – 90%.^[27-29]^ The inter-code variability in accuracy likely depends on multiple factors—perceived disease severity of the assigned healthcare provider, experience of the assigned provider, complexity of the disease-specific ICD coding architecture, decision-support within the local electronic health record, availability of medical coders, and the coding culture within the facility.

As medical coding in the Military Health System is not directly tied to reimbursement, one might expect military ICD coding to be less accurate than other healthcare sectors. The Electronic Surveillance System for the Early Notification of Community-based Epidemics (ESSENCE) uses syndromic groupings from ICD codes for early outbreak detection. From a record review of 2,474 records, investigators found an excellent interrater consistency between 0.87-1.0.^[30]^ While the accuracy fluctuated between diseases, this ESSENCE study demonstrated the efficacy of using ICD codes within a military surveillance system.

ICD-based injury severity measures have also been used to predict in-hospital mortality among injury-related admissions. From a systematic review of 22 eligible studies, the reported area under the receiver operating characteristic curve ranged from 0.681-0.958 indicating a wide range of predictive ability.^[31]^ ICD codes may be used in a predictive fashion, but their accuracy will depend on their employment method.

In ASIMS medical providers attach profiles to ICD codes, but as previously discussed, they assign durations based on calendar convenience almost half the time. This study’s models, rather than predicting physiological duration by diagnosis, might be predicting the profiling behavior of medical providers.

ICD-9-CM codes also comprised 84.6% of the primary diagnostic codes in the original dataset, and all were forward mapped to ICD-10-CM codes. Generally, ICD-9-CM codes are less specific than ICD-10-CM codes; thus, forward mapping inherently results in a loss of diagnostic specificity. Additionally, codes were mapped in a one-to-one fashion, only keeping the ICD-10-CM code with the highest prevalence among profiles published after 30 Sep 2015. This approach also likely contributed to the models’ poor performance.

When a one-to-one match was unavailable in translating AFSCs to job classes, the determination made may have been subject to unconscious bias of the author. For instance, pilots and physicians have obvious civilian correlates; however, matching the AFSC title of Mission Generation Vehicular Equipment Maintenance to a civilian parallel was more difficult.

### Strengths

Ironically, while this study did not produce a tool with much clinical accuracy, it added evidence for the need of such decision support and a potential model for development. If providers are assigning restriction durations based upon calendar convenience, then they are not considering patient-specific characteristics, occupational requirements, and disease recovery timelines. This is concerning and warrants further investigation.

While this is the first study to apply quantile regression to disability data, it also offers an example of the importance of data quality to model development. Even with additional covariates, including prescribed medications, laboratory results, radiological studies, and vital signs, the available profile duration data within ASIMS is likely not of sufficient quality to build a military-specific occupational disposition tool similar to MDGuidelines^®^ (Reed Group, Westminster, CO, USA) and the Official Disability Guide^®^ (MCG Health, Seattle, WA, USA).^[32,33]^ For now military providers should consult civilian-facing tools of this nature when building profiles for active duty service members, heeding attention that they were built with non-representative populations.

## Conclusions

This study failed to produce an accurate clinical tool to estimate occupational disposition durations in AF service members. While medical readiness is the core purpose of the Military Health System,^[1,2]^ and a tool of this nature could improve the validity of readiness statistics and the safety of return-to-work recommendations, a high-quality, representative data source is not available at this time. Tools such MDGuidelines^®^ (Reed Group, Westminster, CO, USA) and the Official Disability Guide^®^ (MCG Health, Seattle, WA, USA) ^[32,33]^ might still be helpful, but the AF should consider occupational medicine training for its non-Flight Surgeon providers and building evidence-based return-to-work protocols. Failing in its primary goal, this study still produced several useful findings warranting further investigation—the indirect correlation between profile duration and very heavy job class and the assignment of profile durations based on convenient calendar times.

## Data Availability

All data referenced in the manuscript are from the Aeromedical Services Information Management System (ASIMS) and provided by the Defense Health Agency (DHA). It is not freely available and requires specialized access and a data use agreement.

